# Perceived preparedness to respond to the COVID-19 pandemic: A study with healthcare workers in Ghana

**DOI:** 10.1101/2020.07.10.20151142

**Authors:** Patience A. Afulani, Akua O. Gyamerah, Raymond Aborigo, Jerry John Nutor, Hawa Malechi, Amos Laar, Mona Sterling, John Koku Awoonor-Williams

## Abstract

**Introduction:** Healthcare workers’ (HCWs) preparedness to respond to pandemics is critical to containing disease spread. Low-resource countries, however, experience barriers to preparedness due to limited resources. In Ghana, a country with a constrained healthcare system and high COVID-19 cases, we examined HCWs’ perceived preparedness to respond to COVID-19 and associated factors.

**Methods:** 472 HCWs completed questions in a cross-sectional self-administered online survey. Perceived preparedness was assessed using a 15-question scale (Cronbach alpha=0.91) and summative scores were created (range=0-45). Higher scores meant greater perceived preparedness. We used linear regression with robust standard errors to examine associations between perceived preparedness and potential predictors.

**Results:** The average preparedness score was 24 (SD=8.9); 27.8% of HCWs felt prepared. In multivariate analysis, factors associated with higher perceived preparedness were: training (β=3.35, 95%CI: to 4.69); having adequate PPE (β=2.27, 95%CI: 0.26 to 4.29), an isolation ward (β=2.74, 95%CI: 1.15 to 4.33), and protocols for screening (β=2.76, 95%CI: 0.95 to 4.58); and good perceived communication from management (β=5.37, 95%CI: 4.03 to 7.90). When added to the model, perceived knowledge decreased the effect of training by 28.0%, although training remained significant, suggesting a partial mediating role. Perceived knowledge was associated with a 6-point increase in perceived preparedness score (β=6.04, 95%CI: 4.19 to 7.90).

**Conclusion:** HCWs reported low perceived preparedness to respond to COVID-19. Training, clear protocols, PPE availability, isolation wards, and communication play an important role in increasing preparedness. Government stakeholders must institute necessary interventions to increase HCWs’ preparedness to respond to the ongoing pandemic and prepare for future pandemics.

**Strengths and limitations of this study:**

- This is one of the few studies globally to empirically examine Healthcare workers’ (HCWs) perceived preparedness to respond to COVID-19, and the first study to do so among HCWs in Ghana and in Africa.
- We developed a perceived preparedness for COVID-19 scale and conducted psychometric analysis to confirm its validity and reliability; this scale can facilitate similar research in other settings.
- We show that providers have low perceived preparedness to respond to the COVID-19 pandemic, and that this is associated with a lack of training on COVID-19, PPE, clear protocols, and isolation wards, as well as poor communication from management.
- The use of an online survey with recruitment via social media may have accounted for a relatively young sample.
- Findings are based on self-reported data from a cross-sectional survey, thus, there may be social desirability bias and associations described are not causal.

## INTRODUCTION

In the aftermath of the Ebola and Zika outbreaks, a 2017 World Bank study reported that countries across the world were inadequately prepared to respond to pandemics.^1^ Despite warnings and subsequent efforts to strengthen global pandemic preparedness, many countries remain underprepared to respond to the novel Coronavirus Disease of 2019 (COVID-19) pandemic due to limited resources, underinvestment, and competing priorities.^1^

As of July 2^nd^, 2020, there were over 10.8 million COVID-19 cases and 520,000 deaths globally.^2^ In all countries, health systems have struggled to procure adequate personal protective equipment (PPE) for healthcare workers (HCWs), testing kits, and hospital beds. Emerging empirical studies report that HCWs have inadequate protocols, knowledge, PPE, and other preparedness indicators to respond to COVID-19.^3–6^

Epidemics and pandemics are often unpredictable by nature. Thus, along with mitigation and suppression strategies, health systems, and in particular, HCWs’ preparedness to respond to pandemics are critical to containing disease spread.^7–10^ Previous studies on epidemics, such as with other Severe Acute Respiratory Syndrome (SARS) and Ebola, have found that preparedness of HCWs are not only essential to effectively containing epidemics, but also in ensuring that they are not pulled from addressing other illnesses that may lead to preventable deaths.^8^

Unfortunately, low-resource countries, like Ghana, experience multilevel barriers to preparedness due to limited resources and weak healthcare infrastructure. Across Africa, weak health systems caused by debt, poor governance and economic instability have made the continent underprepared to contain the spread of COVID-19.^11,12^ For instance, in a 2016 study, African countries reported the lowest scores for preparedness indicators and only two-thirds of countries had a national health emergency preparedness and response plan.^1^ Additionally, a World Health Organization COVID-19 readiness study found that about nine intensive care unit beds are available per one million people across the continent.^13^ Due to challenges like these, the United Nations (UN) has warned of the possible loss of 300,000 to 3.3 million lives in Africa due to COVID-19.^11^

As of July^2nd^, 2020, Ghana had over 18,100 COVID-19 cases and 117 deaths, making it the country with the fourth highest number of cases in Africa and 56^th^ globally.^2^ With less than one hospital bed and 02 physicians per 1,000 people,^11,13^ the country’s constrained health system presents challenges to slowing the spread of the epidemic and in maintaining an overburdened healthcare infrastructure. Yet, few studies have examined HCWs’ preparedness to respond to epidemics in Ghana and none on a pandemic of this scale. Previous studies that assessed HCWs’ preparedness to respond to the Ebola outbreak found that providers did not feel adequately prepared or trained to respond to Ebola, and reported challenges such as inadequate staff and PPE, and delayed reporting of cases.^14–17^ To understand the multilevel barriers to containing the spread of COVID-19 in Ghana, we examined HCWs’ perceived preparedness to respond to the pandemic and the associated contributors, including potential mediating factors.

## METHODS

### Context

Healthcare delivery in Ghana is based on a 3-tier system: 1) the primary level, which is delivered by community-based health planning and services compounds, maternity homes, health centers, and district hospitals; 2) the secondary level, implemented by regional hospitals; and 3) the tertiary level, which is run by specialists at the teaching hospitals. Ninety-three percent of facilities in Ghana are Primary Health Care facilities. There are an estimated 1.8 medical doctors and 42 nurses and midwives per 10,000 population in the country.^18,19^ The Ghana Health Service is tasked with establishing effective mechanisms for disease surveillance, prevention, and control nationally^20^ and is currently leading the country’s COVID-19 response. Since the detection of Ghana’s first case on March 12^th^, 2020, several strategies have been adopted to control the epidemic. Key among them is the “3 Ts approach”—Testing, Tracing, and Treatment. Consequently, more symptomatic cases are being reported resulting in overburdened treatment sites. The exponential increase in cases has led the WHO to declare Ghana as one of the countries with an accelerated increase in the number of COVID-19 cases. HCW deaths due to COVID-19 have sparked threats of strike actions by nurses and doctors in Ghana, raising issues related to the health system and HCW preparedness to contain the virus at all levels of the health system.^21,22^

### Sample and data collection

The data are from a cross-sectional study conducted with HCWs in Ghana between April 17^th^ to May 31^st^, 2020. All healthcare workers (i.e., nurses, physicians, and allied health workers) in Ghana were eligible to participate. Providers were recruited virtually through advertising on social media platforms (WhatsApp, Facebook, and direct messaging), and invited to complete a self-administered online survey through a link in the ad. No incentives were provided, and respondents had the option of skipping questions they did not want to respond to. The sample size is 472 providers who completed all the questions relevant for this analysis, out of a total of 648 who initiated the survey. The survey included questions on demographics, perceived preparedness, as well as other questions relevant to the pandemic response described in the measures section. Providers consented to the study by completing the survey.

**Ethical approval** was obtained from the University of California, San Francisco (UCSF) and the Navrongo Health Research Centre (NHRC).

### Patient and public involvement

Patients and/or the public were not involved in the design, conduct, or reporting, or dissemination plans of this research. HCWs however contributed to the development of the survey questions.

### Measures

#### Dependent variable

The outcome variable in this analysis is perceived preparedness, which was assessed using 15 questions (appendix 1) that captured various aspects of preparedness including personal/self, facility/institutional, and mental/psychological preparedness for prevention, diagnoses, management, and education regarding COVID-19. Each question had the following response options: 0 = Not prepared at all, 1 = A little prepared, 2 = Prepared, 3 = Very prepared, 4 = I don’t know about this, and 5 = Not applicable to my role. These questions were developed by our research team after thorough review of the WHO and CDC COVID-19 preparedness Tools and guidelines for Healthcare Professionals and Facilities, and after soliciting questions from healthcare workers on what they thought was relevant to include to assess preparedness.^23,24^ The questions underwent various revisions and were piloted with 10 HCWs in Ghana. Feedback from the piloting was used to finalize the questions. The 15 questions are combined to create a summative preparedness score for the analysis.

#### Independent variables

The key predictor in this analysis was *training on COVID-19* based on the question “Have you had any training on how to respond to the COVID-19 crises?” with a binary Yes/No response. Other predictors included: *perceived availability of PPE (“*Does your facility have adequate PPE?”); *isolation ward for COVID-19 cases in facility (“*Does your facility have a ward for isolating COVID-19 patients?”); *clear guidelines* (“Have you received guidelines on how to report suspected cases of COVID-19?”, “Does your facility have a protocol for screening for potential COVID-19 patients?”, and “Does your facility have a protocol for managing confirmed COVID-19 patients?”); *communication from management* (“How will you describe communication from management of your facility or your in-charge/supervisor regarding the COVID-19 situation in your facility?”); *ability to isolate at home without exposing family* (“If you have to isolate or quarantine at home because of contact with an infected person, is there a place you can isolate without coming into contact with your family?”); and *perceived knowledge* (“Do you know what to do if you suspect a patient may have COVID-19”, and “Do you know how to manage a confirmed case of COVID-19?”). Provider and facility characteristics are also included as predictors.

#### Analysis

We examined the distribution of variables using descriptive statistics. To create a summative preparedness score, we coded the response options to range from 0 to 3 by recoding “I don’t know about this” (4) to “Not at all prepared” (0), and “Not applicable to my role” (5) to “prepared” (2). Factor analysis of the 15 questions showed they all loaded well on one factor with factor loadings of greater than 0. 3. Cronbach alpha for the 15 items is 0.91. We therefore created a summative score for preparedness ranging from 0-45, where higher scores meant higher perceived preparedness. We categorized scores less than 15 as “Not at all prepared” (equivalent to >1 if divided by number of items (15) to set scale to 0 to 3). Scores 15 to 29 (1 to <2) is considered “somewhat prepared,” and 30 or more (≥ 2) as “prepared”. However, because the continuous preparedness score was normally distributed, we used the continuous score for the analysis presented. We used linear regressions with robust standard errors in bivariate and multivariate analyses to examine the association between perceived preparedness and various predictors. We built multivariate models by gradually adding demographic and other independent variables that were significant in bivariate models and testing model fit and collinearity. Some questions with more than two response options were recoded to binary variables to avoid very small samples in some categories. Variables that did not improve the models or were strongly correlated with other variables were dropped from the final model. Finally, we examined if the relationship between training and preparedness is mediated by perceived knowledge or moderated by type of provider.

#### Sensitivity analysis

Due to the significant number of missing observations (n=166) from people starting and not completing the survey, we compared the characteristics of the analytic sample to the starting sample and ran additional analyses with higher sample sizes by excluding variables that had more missing observations to assess if the main findings changed significantly. Additionally, the survey included general knowledge questions on COVID-19 (e.g., transmission, prevention, symptoms, risk factors, treatment, etc.) which were used to generate a knowledge score. This variable is not included in the current analysis because of the large number of missing observations on that variable which significantly reduced the sample size for the analysis (from 472 to 389). Thus, in the sensitivity analysis, we ran the final models with the knowledge scores and imputed for the missing observations. In addition, we ran the final model as a logistic model, using two binary preparedness variables comparing those who felt prepared (≥ 30) to the others (not at all or somewhat prepared <30) and also those with above median preparedness scores (≥ 23) to those with below the median scores.

## RESULTS

### Descriptive results

The characteristics of respondents are shown in Table 1. Of the 472 respondents used for this analysis, 20% were doctors, 63% nurses (inclusive of midwives and medical/physician assistants) and 17% other professionals, including medical laboratory professionals, disease control officers, nutritionists and other allied healthcare workers. Twenty-six percent worked in teaching hospitals, 59% in other public hospitals, including regional and district hospitals and health centers, and 15% in private facilities. Twenty-three percent were working in the Greater Accra and Ashanti regions (the initial epicenters), another 23% from the Northern region, and the remaining from other regions of the country. The average age of respondents was 34.3 years (SD=6.1), with 8.3 years of experience (SD=5.8). Approximately half of the respondents identified as male and the other half as female.

**Table 1:** Univariate distributions.

| **Table 1: Univariate distributions** |  |  |
| --- | --- | --- |
|  | **No.** | **%** |
| Total | 472 | 100.0 |
| Provider type |  |  |
| Doctor | 94 | 19.9 |
| Nurse/related | 297 | 62.9 |
| Other | 81 | 17.2 |
| Facility type |  |  |
| Teaching hospital | 124 | 26.3 |
| Other government facility | 276 | 58.5 |
| Private/mission facility | 72 | 15.3 |
| Region |  |  |
| Greater Accra/Ashanti | 107 | 22.7 |
| Northern region | 108 | 22.9 |
| Other Northern | 105 | 22.2 |
| other Southern | 152 | 32.2 |
| Years of experience |  |  |
| 5 or less years | 154 | 32.6 |
| 6 to 10 years | 193 | 40.9 |
| More than 10 years | 125 | 26.5 |
| Ages |  |  |
| Less than 30 | 130 | 27.8 |
| 30 to 39 | 260 | 55.7 |
| 40 to 73 | 77 | 16.5 |
| Gender |  |  |
| Male | 238 | 50.4 |
| Female | 234 | 49.6 |
| No. of children |  |  |
| No children | 145 | 31.4 |
| 1 or 2 children | 212 | 45.9 |
| 3 to 6 children | 105 | 22.7 |
| Marital status |  |  |
| Single | 137 | 29.0 |
| Married | 335 | 71.0 |
| Perceived preparedness |  |  |
| Not at all prepared | 74 | 15.7 |
| A little prepared | 267 | 56.6 |
| Prepared | 131 | 27.8 |
| Training on COVID-19 |  |  |
| No | 216 | 45.8 |
| Yes | 256 | 54.2 |
| Facility has adequate PPEs |  |  |
| No | 356 | 75.4 |
| Yes | 32 | 6.8 |
| I don't know | 84 | 17.8 |
| Facility has COVID-19 isolation ward |  |  |
| No | 143 | 30.3 |
| Yes | 315 | 66.7 |
| I don't know | 14 | 3.0 |
| Guidelines on how to report suspected case |  |  |
| No | 91 | 19.3 |
| Yes | 362 | 76.7 |
| I don't know | 19 | 4.0 |
| Facility has protocol for screening for COVID-19 |  |  |
| No | 75 | 15.9 |
| Yes | 378 | 80.1 |
| I don't know | 19 | 4.0 |
| Facility has protocol for managing COVID-19 |  |  |
| No | 168 | 35.6 |
| Yes | 233 | 49.4 |
| I don't know | 71 | 15.0 |
| Communication from management |  |  |
| Very poor communication | 61 | 12.9 |
| Poor communication | 152 | 32.2 |
| Good communication | 218 | 46.2 |
| Very good communication | 41 | 8.7 |
| Ability to isolate at home without exposing family |  |  |
| No | 257 | 54.4 |
| Somewhat | 60 | 12.7 |
| Yes | 155 | 32.8 |
| Know what to do if COVID-19 suspected |  |  |
| No | 23 | 4.9 |
| Somewhat | 134 | 28.4 |
| Yes | 315 | 66.7 |
| Know how to manage a confirmed case of COVID-19 |  |  |
| No | 162 | 34.3 |
| Somewhat | 158 | 33.5 |
| Yes | 103 | 21.8 |
| Not applicable to my role | 49 | 10.4 |

The average score for preparedness was 24 (SD=8.9). Based on specified cut offs, 27.8% felt prepared, 56.6% somewhat prepared, and 15.7% not at all prepared (Table 2). Fifty-four percent had participated in a COVID-19 training and only seven percent reported their facilities had enough PPE. Two-thirds (67%) reported they had an isolation ward for COVID-19 cases in the facility; 76% reported they had guidelines on how to report suspected cases of COVID-19; 80% reported their facility had a protocol for screening for potential COVID-19 patients; and 49% reported they had a protocol for managing confirmed COVID-19 patients. Fifty-five percent perceived communication from management to be good or very good. Only a third (32.8%) were certain of a place to quarantine at home without contact with their family. Two-thirds (66.7%) reported they knew what to do if they suspected a patient may have COVID-19 and only 22% said they know how to manage a confirmed COVID-19 case.

**Table 2:** Bivariate Distributions.

|  | Preparedness scores |  |  |  |  |  |
| --- | --- | --- | --- | --- | --- | --- |
| | N | Mean | Sd | $\beta$ | [95% CI] | |
| Total | 472 | 24.0 | 8.9 |  |  |  |
| Provider type |  |  |  |  |  |  |
| Doctor | 94 | 24.0 | 8.4 | 0.00 | [0 | 0] |
| Nurse/related | 297 | 23.8 | 9.1 | -0.12 | [-2.19 | 1.95] |
| Other | 81 | 24.8 | 8.6 | 0.82 | [-1.83 | 3.48] |
| Facility type |  |  |  |  |  |  |
| Teaching hospital | 124 | 25.1 | 8.7 | 0.00 | [0 | 0] |
| Other government facility | 276 | 23.3 | 9.0 | -1.75 | [-3.64 | 0.13] |
| Private/mission facility | 72 | 24.9 | 8.6 | -0.15 | [-2.73 | 2.43] |
| Region |  |  |  |  |  |  |
| Greater Accra/Ashanti | 107 | 24.4 | 7.7 | 0.00 | [0 | 0] |
| Northern region | 108 | 23.5 | 9.4 | -0.89 | [-3.26 | 1.48] |
| Other Northern | 105 | 22.2 | 8.9 | -2.18 | [-4.57 | 0.20] |
| other Southern | 152 | 25.5 | 9.2 | 1.12 | [-1.08 | 3.31] |
| Years of experience |  |  |  |  |  |  |
| 5 or less years | 154 | 24.3 | 9.7 | 0.00 | [0 | 0] |
| 6 to 10 years | 193 | 23.2 | 8.5 | -1.09 | [-2.98 | 0.80] |
| More than 10 years | 125 | 24.9 | 8.3 | 0.63 | [-1.47 | 2.73] |
| Ages |  |  |  |  |  |  |
| Less than 30 | 130 | 24.1 | 9.5 | 0.00 | [0 | 0] |
| 30 to 39 | 260 | 23.3 | 8.7 | -0.89 | [-2.75 | 0.97] |
| 40 to 73 | 77 | 26.4 | 8.1 | 2.27 | [-0.22 | 4.76] |
| Gender |  |  |  |  |  |  |
| Male | 238 | 25.2 | 8.7 | 0.00 | [0 | 0] |
| Female | 234 | 22.8 | 8.9 | -2.36** | [-3.96 | -0.77] |
| No. of children |  |  |  |  |  |  |
| No children | 145 | 24.3 | 9.2 | 0.00 | [0 | 0] |
| 1 or 2 children | 212 | 23.1 | 8.5 | -1.12 | [-3.01 | 0.77] |
| 3 to 6 children | 105 | 25.2 | 9.5 | 0.95 | [-1.30 | 3.20] |
| Marital status |  |  |  |  |  |  |
| Single | 137 | 25.2 | 8.8 | 0.00 | [0 | 0] |
| Married | 335 | 23.5 | 8.9 | -1.71 | [-3.48 | 0.057] |
| Training on COVID-19 |  |  |  |  |  |  |
| No | 216 | 20.0 | 8.4 | 0.00 | [0 | 0] |
| Yes | 256 | 27.5 | 7.8 | 7.52*** | [6.06 | 8.99] |
| Facility has adequate PPEs |  |  |  |  |  |  |
| No | 356 | 23.4 | 8.7 | 0.00 | [0 | 0] |
| Yes | 32 | 31.1 | 7.0 | 7.66*** | [4.50 | 10.8] |
| I don't know | 84 | 24.0 | 9.2 | 0.62 | [-1.46 | 2.69] |
| Facility has COVID-19 isolation ward |  |  |  |  |  |  |
| No | 143 | 20.3 | 8.2 |  |  |  |
| Yes | 315 | 26.2 | 8.4 | 5.86*** | [4.21 | 7.51] |
| I don't know | 14 | 14.2 | 7.4 | -6.09** | [-10.7 | -1.51] |
| Guidelines on how to report suspected case |  |  |  |  |  |  |
| No | 91 | 18.3 | 8.4 | 0.00 | [0 | 0] |
| Yes | 362 | 25.7 | 8.3 | 7.49*** | [5.57 | 9.42] |
| I don't know | 19 | 19.2 | 8.2 | 0.91 | [-3.24 | 5.05] |
| Facility has protocol for screening for COVID-19 |  |  |  |  |  |  |
| No | 75 | 18.6 | 8.1 | 0.00 | [0 | 0] |
| Yes | 378 | 25.5 | 8.5 | 6.92*** | [4.83 | 9.01] |
| I don't know | 19 | 16.5 | 8.9 | -2.05 | [-6.29 | 2.20] |
| Facility has protocol for managing COVID-19 |  |  |  |  |  |  |
| No | 168 | 20.9 | 8.0 | 0.00 | [0 | 0] |
| Yes | 233 | 26.7 | 8.5 | 5.78*** | [4.09 | 7.47] |
| I don't know | 71 | 22.7 | 9.5 | 1.83 | [-0.54 | 4.19] |
| Communication from management |  |  |  |  |  |  |
| Very poor communication | 61 | 17.7 | 8.0 | 0.00 | [0 | 0] |
| Poor communication | 152 | 20.5 | 8.0 | 2.81* | [0.49 | 5.14] |
| Good communication | 218 | 26.8 | 7.8 | 9.11*** | [6.89 | 11.3] |
| Very good communication | 41 | 32.2 | 6.9 | 14.5*** | [11.4 | 17.6] |
| Ability to isolate at home without exposing family |  |  |  |  |  |  |
| No | 257 | 23.0 | 9.0 | 0.00 | [0 | 0] |
| Somewhat | 60 | 22.3 | 7.9 | -0.66 | [-3.13 | 1.80] |
| Yes | 155 | 26.4 | 8.7 | 3.46*** | [1.72 | 5.21] |
| Know what to do if COVID-19 suspected |  |  |  |  |  |  |
| No | 23 | 14.7 | 7.8 | 0.00 | [0 | 0] |
| Somewhat | 134 | 19.2 | 7.4 | 4.59* | [1.05 | 8.12] |
| Yes | 315 | 26.8 | 8.2 | 12.1*** | [8.72 | 15.5] |
| Know how to manage a confirmed case of COVID-19 |  |  |  |  |  |  |
| No | 162 | 18.9 | 7.9 | 0.00 | [0 | 0] |
| Somewhat | 158 | 24.2 | 8.0 | 5.33*** | [3.61 | 7.04] |
| Yes | 103 | 30.4 | 7.4 | 11.5*** | [9.54 | 13.4] |
| Not applicable to my role | 49 | 27.1 | 7.5 | 8.21*** | [5.71 | 10.7] |

95% confidence intervals in brackets \* p<0.05; \*\* p<0.01; \*\*\* p<0.001

### Bivariate results

In the bivariate analysis (Table 2), significant factors associated with preparedness were male gender; training on COVID-19; availability of PPE, isolation ward, protocols for diagnoses and management; communication from management; ability to isolate at home without exposing family; and confidence in knowledge of what to do for a suspected case and management of COVID-19. For example, the average preparedness score was about 28 (SD=7.8) for those who received training compared to 20 (SD=8.4) for those who had no training; and 31 (SD=7.0) for those who reported their facility had adequate PPE compared to 23 (SD=8.7) for those who reported they did not have enough PPE.

### Multivariate analysis

In the multivariate analysis (Table 3), including only the demographic models (model 1), training was associated with a 7-point higher score in preparedness (β=7.47, 95%CI: 5.94 to 9.01, p<0.001), and decreased to about 5 points (β=4.64, 95%CI: 3.27 to 6.01; p<0.001) with the addition of availability of PPE, isolation ward, and protocols for diagnoses and management; perceived communication from management; and ability to self-isolate at home to the model (model 2). When knowledge of how to manage a COVID-19 patient was added to the model (model 3), the training effect decreased by about 28% (β=3.35, 95%CI: 2.01 to 4.69; p<0.001), suggesting the effect of training is partially mediated by perceived knowledge. Perceived knowledge of how to manage a COVID-19 patient is associated with a 6-point higher perceived preparedness score compared to not knowing what to do (β=6.04, 95%CI: 4.19 to 7.90; p<0.001). In the final model (model 3), having adequate PPE (β=2.27, 95%CI: 0.26 to 4.29; p<0.05), an isolation ward (β=2.74, 95% CI: 1.15 to 4.33; p<0.001), protocols for screening (β=2.76, 95%CI: 0.95 to 4.58; p<0.01), and good perceived communication from management (β=5.37, 95%CI: 4-03 to 7.90; p<0.001) were associated with higher perceived preparedness.

**Table 3:** Multivariable linear regression of potential predictors on Preparedness.

95% confidence intervals in brackets \* p<0.05; \*\* p<0.01; \*\*\* p<0.001
|  |  |  |  |  |  |  |  |  |  |
| --- | --- | --- | --- | --- | --- | --- | --- | --- | --- |
|  | *Model 1* |  |  | *Model 2* |  |  | *Model 3* |  |  |
| | $\beta$ | *[95% CI]* | | $\beta$ | *[95% CI]* | | $\beta$ | *[95% CI]* | |
| Had COVID-19 training | 7.47*** | [5.94 | 9.01] | 4.64*** | [3.27 | 6.01] | 3.35*** | [2.01 | 4.69] |
| Provider type |  |  |  |  |  |  |  |  |  |
| Doctor | 0 | [0 | 0] | 0 | [0 | 0] | 0 | [0 | 0] |
| Nurse/related | 2.85** | [0.71 | 4.99] | 3.70*** | [1.87 | 5.53] | 3.48*** | [1.76 | 5.21] |
| Other | 2.96* | [0.48 | 5.44] | 2.48* | [0.33 | 4.64] | 2.26* | [0.077 | 4.44] |
| Region |  |  |  |  |  |  |  |  |  |
| Teaching hospital | 0 | [0 | 0] | 0 | [0 | 0] | 0 | [0 | 0] |
| Other government facility | -3.15** | [-5.32 | -0.99] | -2.03 | [-4.07 | 0.0064] | -1.75 | [-3.65 | 0.16] |
| Private/mission facility | -2.93* | [5.67 | -0.20] | -2.87* | [-5.30 | -0.44] | -2.71* | [-5.01 | -0.41] |
| Region |  |  |  |  |  |  |  |  |  |
| Greater Accra/Ashanti | 0 | [0 | 0] | 0 | [0 | 0] | 0 | [0 | 0] |
| Northern region | -3.48** | [-5.94 | -1.01] | -2.69* | [-4.87 | -0.52] | -2.29* | [-4.41 | -0.17] |
| Other Northern | -4.09** | [-6.56 | -1.62] | -3.53** | [-5.71 | -1.36] | -2.69* | [-4.83 | -0.55] |
| other Southern | -1.5 | [-3.75 | 0.75] | -1.96 | [-3.94 | 0.016] | -1.97* | [-3.92 | -0.024] |
| Years of experience |  |  |  |  |  |  |  |  |  |
| 5 or less years | 0 | [0 | 0] | 0 | [0 | 0] | 0 | [0 | 0] |
| 6 to 10 years | -0.66 | [-2.38 | 1.06] | -0.95 | [-2.45 | 0.55] | -0.9 | [-2.32 | 0.53] |
| More than 10 years | 0.8 | [-1.15 | 2.75] | 0.24 | [-1.47 | 1.95] | 0.16 | [-1.50 | 1.82] |
| Female | -1.46 | [-2.99 | 0.076] | -1.88** | [-3.24 | -0.53] | -1.43* | [-2.71 | -0.15] |
| PPEs Adequate |  |  |  | 2.14* | [0.064 | 4.21] | 2.27* | [0.26 | 4.29] |
| Have Isolation ward |  |  |  | 3.53*** | [1.94 | 5.12] | 2.74*** | [1.15 | 4.33] |
| Protocol for screening for COVID-19 |  |  |  | 2.98** | [1.17 | 4.79] | 2.76** | [0.95 | 4.58] |
| Good communication from management |  |  |  | 6.01*** | [4.67 | 7.34] | 5.37*** | [4.03 | 6.72] |
| Place to isolate at home |  |  |  | 1.27 | [-0.12 | 2.67] | 1.06 | [-0.30 | 2.41] |
| Know how to manage COVID-19 case |  |  |  |  |  |  |  |  |  |
| Somewhat |  |  |  |  |  |  | 2.84*** | [1.27 | 4.42] |
| Yes |  |  |  |  |  |  | 6.04*** | [4.19 | 7.90] |
| Not applicable to my role |  |  |  |  |  |  | 4.19*** | [2.07 | 6.31] |
| Constant | 22.9*** | [20.3 | 25.6] | 15.1*** | [12.1 | 18.1] | 13.7*** | [10.8 | 16.6] |
| N | 472 |  |  | 472 |  |  | 472 |  |  |
| R-squared | 0.223 |  |  | 0.427 |  |  | 0.477 |  |  |

Other predictors in the final model were provider and facility type, region, and gender. Nurses and other non-physician providers had higher preparedness scores than doctors. But, the interaction between provider type and training was not significant, suggesting the relationship between training and preparedness does not differ between providers. Also, providers in private facilities had lower preparedness scores than those in teaching hospitals, but there was no significant difference between providers in teaching hospitals and other government facilities. In addition, providers in all the other regions had lower preparedness scores than providers in Greater Accra and Ashanti region, which were the initial and current epicenters; and female providers had lower perceived preparedness than males.

### Sensitivity Results

The analytic sample did not differ substantially from the total sample on key variables, except that there were more providers from teaching hospitals in the analytic sample (26%) compared to the total sample (15%). Also, the results obtained in the various sensitivity analyses (appendix 2) did not differ substantially from the results presented. The average knowledge score was 53 (SD=4.19) out of 66 for the 392 providers who responded to the knowledge questions. But knowledge scores were not associated with perceived preparedness (using the continuous knowledge variable, as well as a categorical variable and imputing for the missing observations). Other findings were consistent in their significance, direction, and magnitude of the associations. Providers who had participated in COVID-19 related training had over two times higher odds of being prepared compared to those who had no training.

## DISCUSSION

Our study presents evidence on perceived preparedness to respond to the COVID-19 pandemic among healthcare workers in Ghana. Based on a perceived preparedness for COVID-19 scale, we found that most HCWs do not feel prepared to respond to the pandemic. Low perceived preparedness was associated with lack of training, PPE, clear protocols, and isolation wards as well as poor communication from management. The effect of training was partially mediated by perceived knowledge. This study is one of the few studies to empirically examine providers’ perceived preparedness for COVID-19, and the first to use the perceived preparedness scale, which was developed by our study team. The process of developing the questions ensured that the scale had high content validity, and the psychometric analysis showed it has high construct validity and reliability. The association of perceived preparedness with various factors in theoretically predictable ways also provides evidence of criterion validity. The scale, thus, has good psychometric properties with potential utility for replication in other settings.

Our findings are consistent with what we expected given that all the predictors are critical to preventing the spread of COVID-19 and for management and containment. They are also consistent with findings from the few emerging studies on provider preparedness to respond to COVID-19 elsewhere. For example, a study in Palestine found that the vast majority of HCWs did not have access to masks and other PPE and only 11.6% felt prepared to respond to the epidemic.^3^ Another study in Jordan found that about half of medical doctors surveyed had access to an institutional COVID-19 protocol and a minority had PPE.^4^ As in our study, the doctors who reported having an institutional protocol for dealing with COVID-19 cases and those who reported sustained availability of PPE had higher preparedness scores than their references. Also, as in prior studies, males had higher perceived preparedness scores than females.^4^ Another study on maternal and newborn health professionals found that only one-third of respondents received COVID-19 training.^6^ Moreover, similar to our finding that 49% of HCWs reported receiving a protocol for COVID-19 care provision, only half of the providers in low-middle income countries received updated care provision guidelines compared to 82% of those in high income countries.

The finding that the effect of training on preparedness is partially accounted for by perceived knowledge to manage cases is likely because perceived knowledge increases self-efficacy. However, general knowledge scores were not associated with preparedness in the sensitivity analysis, which is potentially due to the nature of the knowledge questions, the high knowledge scores in the sample, and the extent of missingness on that variable. Studies in China and Vietnam also found that HCWs surveyed had good knowledge about COVID-19 transmission, signs, symptoms, and prevention.^25,26^ However, studies in Pakistan and the United Arab Emirates found that a majority of HCWs surveyed had poor knowledge of COVID-19.^5,27^ The relatively high knowledge among HCWs in Ghana might be due to the increase of education as more is learnt about the disease.

### Implications in the context of the health system in Ghana and other low-resource settings

The UN estimates that up to 3.3 million people in Africa could die of COVID-19 if containment measures are not prioritized.^11^ With fragmented health systems that were already constrained before the advent of COVID-19, low- and middle-income countries, such as Ghana, must act differently to avoid such a calamity. Healthcare workers are central to containment efforts and the results of our study suggest that most of them are not prepared to respond to the pandemic. Ghana has received support from the International Monetary Fund and the World Bank, and other sources to help with containment efforts.^28,29^ Prioritizing the use of these funds to make HCWs more prepared to effectively respond to COVID-19 cases is key to the country’s containment efforts. This should include the provision of adequate PPE, training on protocols for screening, diagnoses and management of cases and providing clear care guidelines. The approach should be comprehensive and inclusive of HCWs in the private sector, since current efforts largely focus on only geographic hotspots and government facilities.

### Limitations and Strengths

A key limitation of this study is the sampling approach. Specifically, the use of an online survey with recruitment via social media may have accounted for the relatively young sample. Thus, this sampling limitation and volunteer bias will affect the generalizability of the findings to all HCWs in Ghana. Nonetheless, this is the first study to our knowledge assessing HCWs preparedness for COVID-19 in Ghana and in Africa, thus, study findings are instructive to the current pandemic response. In addition, given that the country was on partial lock-down during the study period, an online survey was the best option available. A second limitation is social desirability bias from the self-reported data—providers may want to project a greater sense of preparedness than they actually have. The use of composite scores from several questions helps address this limitation. Finally, because this was a cross-sectional study, associations described are not causal.

## CONCLUSIONS

We found that HCWs had low perceived preparedness to respond to COVID-19. Training, clear protocols, PPE availability, isolation wards, and good communication from management play an important role in increasing provider preparedness. Given the devastating implications of low preparedness in response to the pandemic in Africa as warned by the UN, it is critical for the government of Ghana and other stakeholders within the health system to intervene to increase HCWs’ preparedness to respond to the rapidly growing epidemic in the country. In addition, the assessment tool developed for this work provides an opportunity to evaluate preparedness of healthcare workers in other settings to inform the global response to the pandemic. Importantly, this evaluation will also provide a baseline of HCW preparedness that can be tracked over time to further address the barriers in the health systems faced by providers globally.

## Data Availability

Study data are available on request. All relevant data are included in the article or enclosed as supplementary information.

## Acknowledgements

We wish to thank all healthcare providers who participated in the study and who helped in the survey dissemination.

## Contributor

PA conceptualized study, developed data collection instruments, conducted data analysis, and led manuscript writing; AG helped develop data collection instruments and write manuscript; RA helped develop data collection instruments and write manuscript; JN helped develop data collection instruments and write manuscript; HM helped develop data collection instruments and write manuscript; AL helped develop data collection instruments and write manuscript; MS helped develop data collection instruments and write manuscript; JA helped develop data collection instruments and write manuscript. All authors read and approved final version.

## Patient consent for publication

Not required.

## Ethics approval

The proposal and materials for the study that provides data for this manuscript were approved by the ethical review units of the University of California, San Francisco and Navrongo Health Research Center in Ghana.

## Provenance and peer review

Not commissioned; externally peer reviewed.

